# Association of Free-Living Diet Composition and Plasma Lipoprotein(a) Levels in Healthy Adults

**DOI:** 10.1101/2023.03.24.23287725

**Authors:** Anastasiya Matveyenko, Heather Seid, Kyungyeon Kim, Nelsa Matienzo, Rajasekhar Ramakrishnan, Tiffany Thomas, Gissette Reyes-Soffer

## Abstract

**Background:** Lipoprotein (a) [Lp(a)] is an apoB100-containing lipoprotein with high levels positively associated with atherosclerotic cardiovascular disease (ASCVD). Lp(a) levels are largely genetically determined. Currently, the only approved therapy for patients with extreme elevations of Lp(a) is lipoprotein apheresis, which eliminates apoB100-containing particles including Lp(a). The current study analyzed the association of free-living diet composition with plasma Lp(a) levels.

**Methods:** Dietary composition data from 28 diverse participants was collected via a standardized protocol by registered dietitians using 24-hour recalls. Data were analyzed with the Nutrition Data System for Research (Version 2018). Diet quality was calculated using the Healthy Eating Index (HEI) score. Fasting plasma Lp(a) levels were measured via an isoform-independent ELISA.

**Results:** Subjects self-reported race/ethnicity [Black (n=18); Hispanic (n=7); White (n=3)]. The mean age was 48.3±12.5 years with 17 males. Median level of Lp(a) was 79.9 nmol/L (34.4-146.0) and was negatively associated with absolute (g/d) and relative (percent calories) intake of dietary saturated fatty acid (SFA) (SFA absolute: R= -0.43, p= 0.02, SFA calorie %: R= -0.38, p= 0.04), absolute palmitic acid intake (palmitic absolute: R= -0.38, p= 0.04), and absolute steric acid intake (steric absolute: R= -0.40, p= 0.03). Analyses of associations with HEI when stratified based on Lp(a) levels > or ≤ 100nmol/L revealed no significant associations with any of the constituent factors.

**Conclusions:** We found a negative relationship between dietary saturated fatty acid intake and Lp(a) levels in a diverse cohort of individuals. The mechanisms underlying this relationship require further investigation.

## INTRODUCTION

Atherosclerotic cardiovascular disease (ASCVD) is the leading cause of death in the United States ^1^. One independent and causal risk factor for developing ASCVD is high plasma level of lipoprotein(a) [Lp(a)] ^2-4^. Lp(a) has two main protein components: an integral membrane protein, apolipoprotein (apo) B100, covalently bound to the glycoprotein apolipoprotein(a) [apo(a)] ^2-4^. Plasma Lp(a) levels are 70-90% determined by the *LPA* gene ^5-7^. Apo(a) varies in size from 300 to 800 kDa due to different numbers of Kringle 4 type 2 (KIV-2) repeats, ranging from 1 to >40. A universal consensus for the threshold of elevated Lp(a) associated with ASCVD risk has not been determined ^8^, hence there are multiple different published cut-off ranges. However, a continuous causal association between Lp(a) and ASCVD is well established ^9^.

Lifestyle modifications, including exercise and diet interventions, are low-cost and effective ways to prevent and help treat cardiovascular disease. Lp(a) levels do not change or may slightly increase (10-15%) after intense exercise training in previously sedentary individuals ^10, 11^. Additionally, unlike other apoB-containing lipoproteins and CVD risk factors (i.e. obesity, insulin resistance), in which diet modifications contribute to a decreased risk of events ^12^, Lp(a) levels do not change during cardio-beneficial diet interventions ^13, 14^. Several studies have examined the possible effects of dietary interventions on Lp(a) ^15-18^. Studies by Ginsberg et al. ^18^, Shin et al. ^15^, and Silaste et al. ^16^ observed a negative relationship between plasma Lp(a) levels and saturated fatty acids (SFA) ^15, 16, 18^. Conversely, Haring et al. found a positive relationship between plasma Lp(a) levels and unsaturated fatty acids ^17^. The studies suggest that overall diet composition may influence Lp(a) levels and may not be in line with diets that provide cardiovascular benefits. To date, none of the studies directly evaluate the relationship between the participants’ free-living diet and Lp(a) levels prior to intervention, which may or may not have contributed to the results on saturated fat and Lp(a) levels observed. Additionally, these studies did not include apo(a) isoform size and race/ethnicity, both known to affect Lp(a) levels ^19^. Therefore, we examined food records from a diverse cohort of subjects previously enrolled for studies that evaluated lipid and lipoprotein metabolism. We evaluated the relationship of Lp(a) levels with diet composition, and diet quality as measured by the Healthy Eating Index (HEI).

## METHODS

### Study Participants

All studies were approved by the CUIMC institutional review board (IRB), and informed consent was obtained from all participants. Participants were healthy volunteers with no history of cardiovascular disease (CVD) or type 2 diabetes (T2D) and did not report taking any lipid-lowering medications ^20, 21^. Dietary data were obtained from screening visits for enrollment in previously completed lipid and lipoprotein metabolism studies at Columbia University Irving Medical Center (CUIMC). Only individuals with complete dietary records were included in the present analysis ^20, 21^.

### Study Procedures

All participants provided signed informed consent before any procedures were performed. Participants were screened at our research center facilities after a 12-hour (hr.) overnight fast. We recorded self-reported race/ethnicity (SRRE). Height and weight were measured using a scale, while wearing a hospital gown and no shoes. These measurements were used to calculate body mass index (BMI). Registered dietitians completed dietary 24-hour recalls, in person. Participants were excluded from this study if they followed non-conventional dietary habits such as the ketogenic diet or intermittent fasting. One dietary recall was obtained per participant via the multiple pass method ^22, 23^. Dietary intake data were analyzed using Nutrition Data System for Research (NDSR) software Version 49 (2018) developed by the Nutrition Coordinating Center (NCC), University of Minnesota, Minneapolis, MN ^24^. Diet macronutrient data were evaluated and included: carbohydrate, protein, and fat, including saturated (SFAs), mono- (MUFAs) and poly- (PUFAs) unsaturated fatty acids as well as dietary fiber (soluble and insoluble). Diet composition was analyzed on an absolute (g/d) and relative basis (% Cal). This observational study examined the relationship of fasting Lp(a) levels with free-living diet composition, particularly fat intake.

### Healthy Eating Index

Microsoft Excel was used to calculate the Healthy Eating Index (HEI) Score 2015 for each participant. HEI-2015 describes the diet quality according to the recommendations outlined in the 2015-2020 Dietary Guidelines for Americans by generating a score from 0 to 100 (100 being 100% in congruence with the guidelines) ^25^. The score is a composite of thirteen factors representing different food groups classically associated (positively or negatively) with chronic disease. The relationship between HEI and Lp(a) level was evaluated in all subjects and in subgroups stratified by Lp(a) level. There is no clinically accepted level to denote elevated Lp(a). We, therefore, stratified our cohort into “high” and “low” using a cut point that considers two published recommendations for Lp(a) levels (“high” - >100nmol/L; “low” - ≤ 100nmol/L). The National Heart, Lung, and Blood Institute (NHLBI) Working Group Recommendations use Lp(a) levels >75nmol/L as “high” ^26^, and the 2019 European Society of Cardiology (ESC)/European Atherosclerosis Society (EAS) Guidelines considers elevated Lp(a) levels as Lp(a) ≥ 125 nmol/L ^27^.

### Laboratory Measurements

Each participant had a 12-hour fasting blood draw via an intravenous (IV) catheter from forearm veins. Briefly, blood was obtained in EDTA containing test tubes, immediately placed on ice, and spun in a centrifuge at 1693 RCF, 4° Celsius (C) for 20 minutes. Plasma was isolated from the test tube and stored in a -80° C freezer. Frozen samples were shipped on dry ice to the laboratory of Dr. Santica Marcovina (Seattle, Washington), where plasma Lp(a) levels were measured using an isoform-independent, double monoclonal antibody-based ELISA assay ^28-30^. In this cohort, Lp(a) levels were not normally distributed, so we calculated medians and interquartile ranges (IQR). Apo(a) isoform size measurements were performed by the same laboratory ^31^. Plasma lipids (total cholesterol (TC), triglycerides (TG), and high-density lipoprotein (HDL) cholesterol) were measured by Integra400plus (Roche). Plasma low-density lipoprotein (LDL) cholesterol levels were estimated using the Friedewald formula. A human enzyme-linked immunosorbent assay measured plasma apoB100 (ELISA) with kit # 3715-1HP-2 from Mabtech, Inc, Cincinnati, OH.

### Weighted Isoform Size Calculations (wIS)

Most individuals express two apo(a) isoforms in plasma, which are generally inversely correlated with Lp(a) plasma levels, with smaller isoforms typically dominating. To account for the difference in percent expression of each isoform, we calculated a weighted isoform size ^32^. Example: If the two allele sizes are 20 and 30, with relative expression of 70% and 30%, respectively, the *wIS* is 0.7*20+0.3*30 = 23.

### Statistical Analysis

Based on previously identified relationships of diet components with lipids, twenty-three dietary variables were identified a priori for analysis from the 170 variables available via NDS-R output. Diet data are presented as absolute [grams/day (g/d)] and relative [percent of total Calories (% Cal)] intake. Pearson correlation and linear regression were used to evaluate relationships between variables using the R software ^33^. Statistical significance was set at a p-value less than 0.05.

## RESULTS

We analyzed diet records from 28 participants, baseline characteristics including lipid and lipoprotein levels are listed in Table 1. The mean age of the cohort was 48.3 ± 12.5 years; 17 out of the 28 subjects were male. The participants were overweight with a mean BMI of 29.5 ± 3.3 kg/m^2^. Eighteen study participants listed Black as their SRRE. Plasma lipid levels (TC, TG, HDL, LDL) and apoB100 levels were within normal ranges. The median Lp(a) level was 79.9 nmol/L (IQR 34.4 - 146 nmol/L) and the calculated *wIS* was 22.4. As observed in larger published cohorts, apo(a) isoform size was negatively associated with Lp(a) levels (Supplemental Figure 1). Individual Lp(a) levels, isoforms size expression and calculated *wIS* for the full cohort are included in Supplemental Table 1.

**Table 1.** Participant Characteristics (N=28)

|  |  |
| --- | --- |
| **Sex (n)** |  |
| *Male* | 17 (60.7) |
| **Race/Ethnicity (n)** |  |
| *Black* | 18 (64.3) |
| *Hispanic* | 7 (25.0) |
| *White* | 3 (10.7) |
| **Age (years)** | 48.3±12.5 |
| **BMI (kg/m<sup>2</sup>)** | 29.5±3.3 |
| **Total Cholesterol (mg/dL)** | 152.8±22.3 |
| **Total Triglyceride (mg/dL)** | 98.5±43.6 |
| **LDL-C (mg/dL)** | 86.6±18 |
| **HDL-C (mg/dL)** | 46.6±12.8 |
| **ApoB100 (mg/dL)** | 90.7±27.2 |
| **Lp(a) (nmol/L)** | 79.9 (34.4-146.0) |
| **Apo(a) *wIS*** | 22.4±4.6 |
| Data are presented as mean ± SD, median (IQR), or n (%). BMI, Body Mass Index; LDL-C, Low-Density Lipoprotein Cholesterol; HDL-C, High-Density Lipoprotein Cholesterol; ApoB100, Apoprotein (B100); Lp(a), Lipoprotein (a); Apo(a), Apoprotein (a); n, number; kg, kilogram; m<sup>2</sup>, meter squared; mg, milligrams; dL, deciliter; IQR, Interquartile Range; SD, Standard Deviation |  |

### Relationships Between Dietary Components plasma lipids, lipoproteins and Lp(a)

ApoB100 levels were positively correlated with total dietary fat (g/d; R = 0.40, p = 0.036), percent Calories from fat (R = 0.52, p = 0.005), total dietary SFA (R = 0.58, p = 0.001), and percent Calories from SFA (R = 0.62, p < 0.001) (Supplemental Table 2).

The relationships between Lp(a) levels and various dietary components were assessed and are presented in Table 2. Apo(a) isoforms and SRRE are determinants of plasma Lp(a) levels ^19^. Our small study size does not allow us to examine independent effects of isoforms and race, due to small cohort, however, we control for these in our linear regression models (Table 2).

**Table 2.** Relationship Between Dietary Variables and Lp(a)

| Dietary Variables |  | Absolute Intake |  |  | Relative Intake |  |  | Absolute Intake† |  | Relative Intake† |  |
| --- | --- | --- | --- | --- | --- | --- | --- | --- | --- | --- | --- |
|  |  | Mean ± SD | R | p-value | Mean ± SD | R | p-value | R<sup>2</sup> | p-value | R<sup>2</sup> | p-value |
|  |  | g/d |  |  | % Cal |  |  |  |  |  |  |
| **Total Energy** | **kcal/d** | 1722.6±599.8 | -0.32 | 0.1 | NA | NA | NA | 0.46 | 0.203 | NA | NA |
| **Macronutrients** | Carbohydrate | 221.8±87.7 | -0.18 | 0.35 | 50.7±12.5 | 0.24 | 0.22 | 0.42 | 0.617 | 0.5 | 0.07 |
|  | Protein | 79.5±35.9 | -0.27 | 0.17 | 18.5±5.8 | 0.03 | 0.90 | 0.47 | 0.156 | 0.42 | 0.66 |
|  | Fat | 60.5±29.9 | -0.34 | 0.08 | 30.5±11.7 | -0.29 | 0.13 | 0.48 | 0.108 | 0.51 | 0.05 |
| **Saturated Fatty Acids** | Total SFA | 18.1±10.8 | -0.43 | 0.02* | 9.2±4.9 | -0.38 | 0.04* | 0.52 | 0.04* | 0.53 | 0.03* |
|  | Palmitic Acid | 10.6±6 | -0.38 | 0.05* | 7.7±4.1 | -0.33 | 0.08 | 0.52 | 0.047* | 0.53 | 0.03* |
|  | Stearic Acid | 4.4±3.1 | -0.4 | 0.03* | 2.3±1.4 | -0.37 | 0.06 | 0.52 | 0.036* | 0.54 | 0.02* |
| **Unsaturated Fatty Acids** | Total MUFA | 22.8±13.1 | -0.14 | 0.47 | 11.4±5.5 | -0.09 | 0.66 | 0.43 | 0.529 | 0.43 | 0.54 |
|  | Palmitoleic Acid | 1.2±0.9 | -0.26 | 0.18 | 0.6±0.5 | -0.17 | 0.4 | 0.44 | 0.308 | 0.43 | 0.44 |
|  | Oleic Acid | 20.9±12.2 | -0.13 | 0.51 | 10.6±5.1 | -0.08 | 0.7 | 0.43 | 0.55 | 0.43 | 0.56 |
|  | Total PUFA | 14.1±8.9 | -0.26 | 0.18 | 7.1±3.3 | -0.18 | 0.37 | 0.47 | 0.152 | 0.47 | 0.14 |
|  | Linoleic Acid | 12.0±8.2 | -0.25 | 0.20 | 6.1±3.1 | -0.18 | 0.37 | 0.47 | 0.149 | 0.47 | 0.13 |
|  | Linolenic Acid | 1.4±0.7 | -0.37 | 0.05 | 0.71±0.3 | -0.25 | 0.19 | 0.49 | 0.092 | 0.46 | 0.19 |
| **Dietary Fiber** | Soluble Fiber | 6.3±4.2 | -0.08 | 0.67 | NA | NA | NA | 0.43 | 0.466 | NA | NA |
|  | Insoluble Fiber | 15.4±11.3 | -0.09 | 0.64 | NA | NA | NA | 0.43 | 0.48 | NA | NA |

SD; Standard deviation. Pearson correlation was used to evaluate the relationships between variables. d, day; g, grams; Kcal, kilocalories; NA, Not Applicable†Linear regression was used to evaluate the relationships between variables, while controlling for wIS and SRRE. wIS, weighted Isoform Size; SRRE, Self-Reported Race/Ethnicity

#### Macronutrients

The mean intakes of total carbohydrate, protein, and fat in our participants were 221.8 ± 87.7g, 79.5 ± 35.9g, and 60.5 ± 29.9g, respectively, corresponding to a macronutrient distribution of 50.7 ± 12.5 % Cal from carbohydrates, 18.5 ± 5.8 % Cal from protein, and 30.5 ± 11.7 % Cal from fat (Table 2). There is no statistically significant relationship between average energy intake (kcal/d) and Lp(a) concentration (R = -0.32, p = 0.10). Additionally, there was a trend towards a negative relationship between fat intake (g/d) and Lp(a) levels (p=0.08), but this relationship did not persist when normalized to total Calories (p=0.13). There were no significant relationships between plasma Lp(a) levels and carbohydrate or protein when compared on an absolute or relative intake (Table 2). However, when we included *wIS* and SRRE in our model the relationships between Lp(a) and relative intake of carbohydrates (p=0.07) and fat (p=0.05) improved (Table 2).

#### Saturated Fatty Acids (SFA) - Palmitic Acid and Stearic Acid

The mean intakes of SFA, palmitic acid, and stearic acid intake were 18.1 ± 10.8g/d (9.2 ± 4.9% Cal), 10.6 ± 6g/d (7.7 ± 4.1% Cal), and 4.4 ± 3.1g/d (2.3 ± 1.4% Cal), respectively (Table 2). There was an inverse relationship between Lp(a) levels with dietary SFA [absolute (R= -0.43, p=0.02) and relative (R=-0.38, p=0.04)], dietary palmitic acid [absolute (R= -0.38, p=0.045)], and dietary stearic acid [absolute (R= -0.4, p=0.034)]. We observed trends toward a negative correlation between Lp(a) and relative intake of palmitic (R= -0.33, p=0.082) and stearic acid (R= -0.37, p=0.056). After controlling for *wIS* and SRRE, we observed an inverse relationship between Lp(a) level and dietary SFA [absolute (R^2^ = 0.52, p=0.04) and relative (R^2^ = 0.53, p=0.28)], palmitic acid [absolute (R^2^ = 0.52, p=0.05) and relative (R^2^ = 0.53, p=0.03)], and stearic acid [absolute (R^2^ = 0.52, p=0.04) and relative (R^2^ = 0.54, p=0.02)] (Table 2). For every one percent increase in calories from SFA, Lp(a) level decreases by 5.974 nmol/L.

#### Unsaturated Fatty Acids - Monounsaturated Fatty Acids (MUFA) and Polyunsaturated Fatty Acids (PUFA)

The mean total MUFA, palmitoleic acid, and oleic acid intake was 22.8 ± 13.1g/d (11.4 ± 5.5% Cal), 1.2 ± 0.9g/d (0.6 ± 0.5% Cal), and 20.9 ± 12.2g/d (10.6 ± 5.1% Cal), respectively (Table 2). The average daily PUFA, linoleic acid, and linolenic acid intake were 14.1 ± 8.9g/d (7.1 ± 3.3% Cal), 12 ± 8.2g/d (6.1± 3.1% Cal), 1.4 ± 0.7g (0.71± 0.3% Cal), respectively. The relationship between plasma Lp(a) and absolute (g/d) and relative (% Cal) MUFA or PUFA intake was not significant, unadjusted, or adjusted for *wIS* and SRRE.

#### Dietary Fiber

The mean soluble and insoluble fiber intake was 6.3 ± 4.2g/d and 15.4 ± 11.3g/d, respectively, and there was no significant relationship with Lp(a).

### Relationships of Lp(a) with Healthy Eating Index

Our cohort had an average HEI score of 57.1± 16, this is similar to results published by NHANES, which found the average HEI score for Americans is 58 ^34^, suggesting that our sample reflects dietary patterns previously described throughout the US population. An HEI score of 100 would suggest perfect alignment with the dietary guidelines. We investigated the relationship between our cohort’s HEI index score and Lp(a) levels and found no relationship between HEI score and Lp(a) level (Figure 2A). When stratified based on high versus low Lp(a) level, the p-value was 0.09, which did not reach statistical significance between HEI and Lp(a) groups (Table 3). Further, when analyzed for each of the thirteen dietary subgroups that make up the HEI sore by unpaired t-test between low and high Lp(a) (Figure 2B), only dietary saturated fat reached statistical significance (p=0.03).

**Table 3:** Relationship between HEI score and Lp(a) value stratified by normal and high Lp(a) levels (N=28)

|  | HEI Score<br>Mean ± SD | p-value |
| --- | --- | --- |
| **Lp(a) Value-Low**<br>≤100 nmol/L (n=15) | 52.4±16 | 0.09 |
| **Lp(a) Value-High**<br>>100 nmol/L (n=13) | 62.6±14.2 |  |
| Legend: Unpaired t-test was used to evaluate the relationships between variables<br>HEI, Healthy Eating Index; SD, Standard Deviation. Alpha significance set at p<0.05 |  |  |

**Figure 1.**
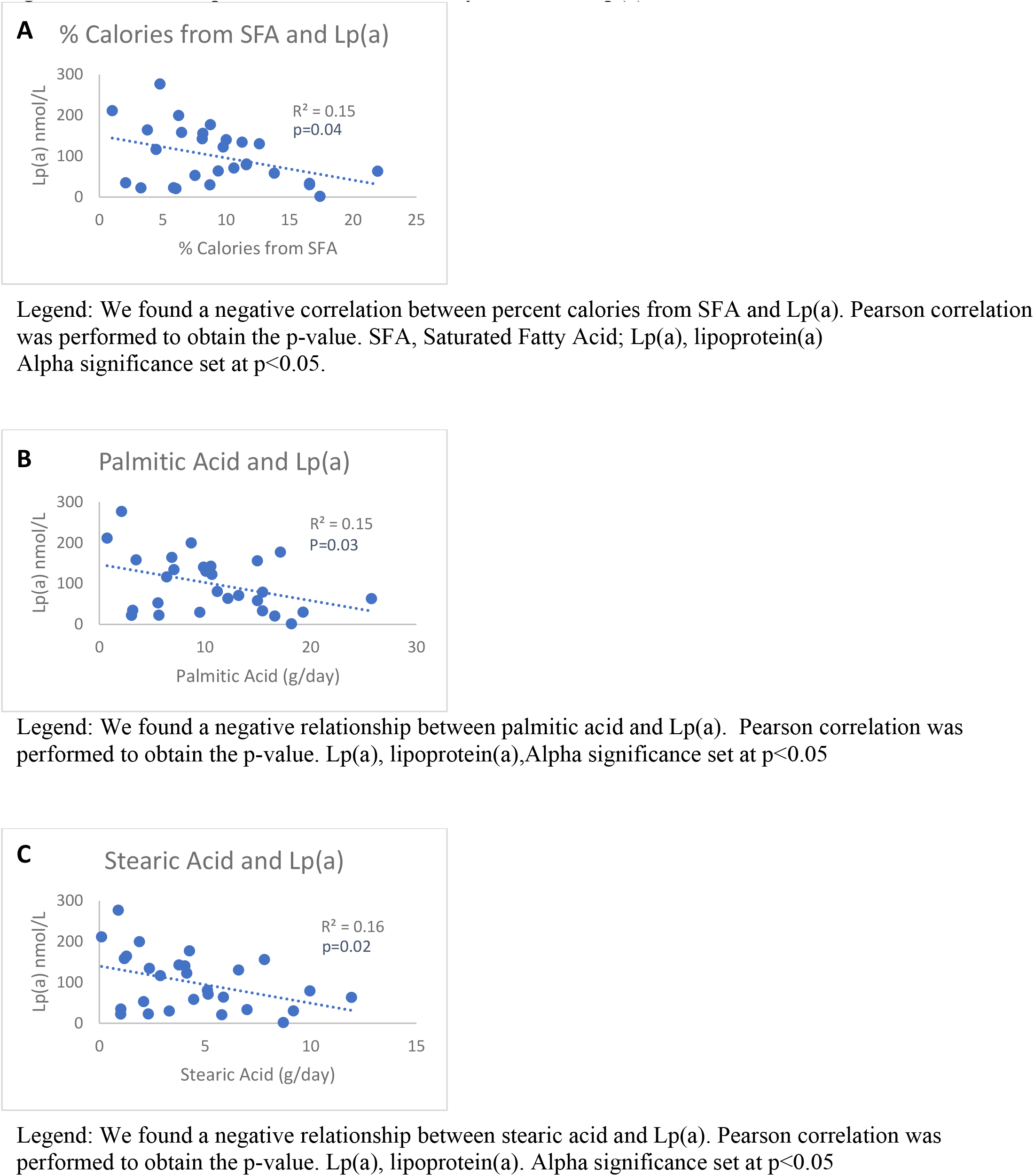
Relationship between Saturated Fatty Acids and Lp(a)

**Figure 2.**
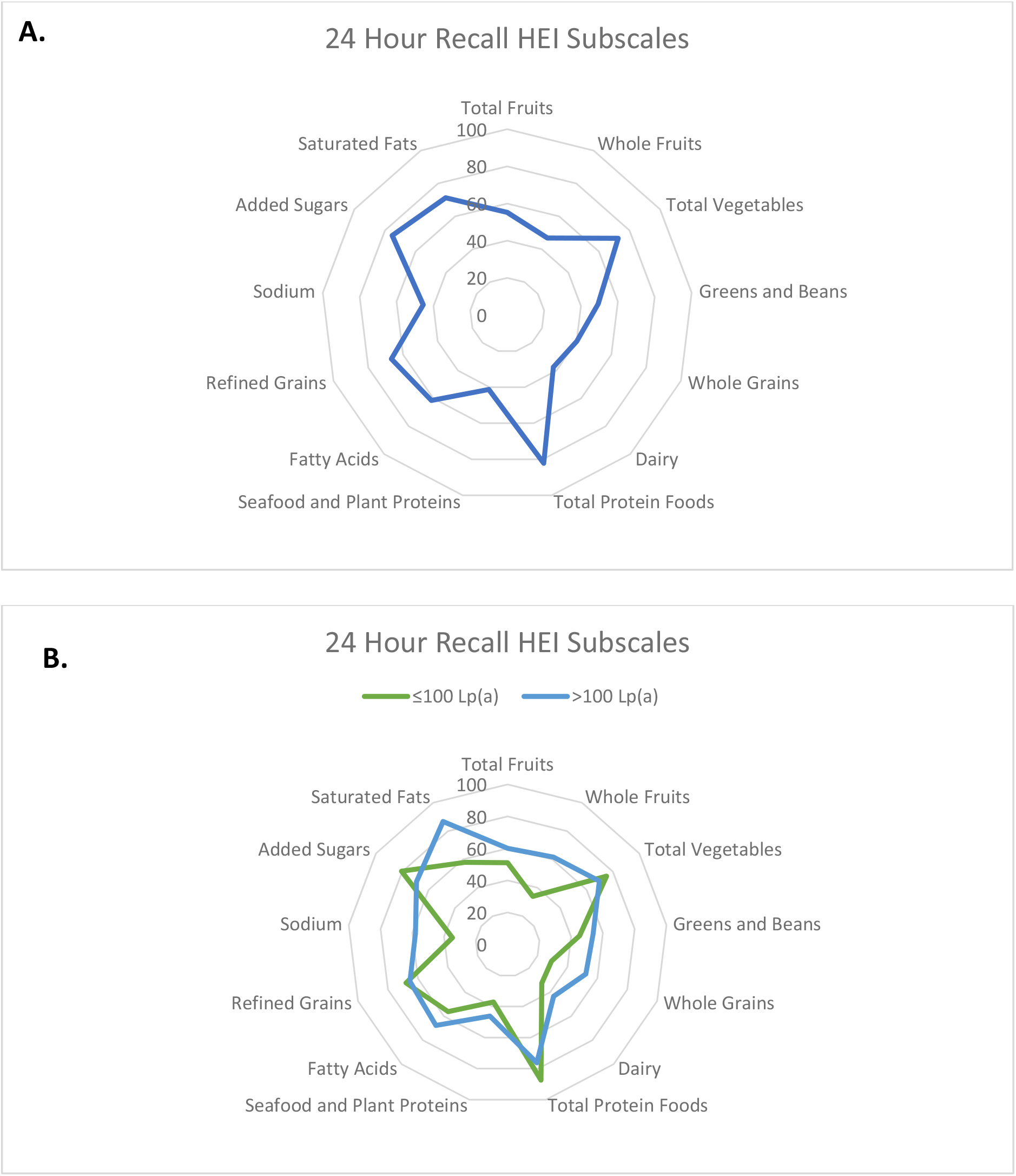
Radar Plots for Healthy Eating Index Broken Down by Food Groups. **A**. Overall HEI assessment based on individual food groups that make up HEI score for all study participants. **B**. We performed unpaired t-test, presenting the relationships between high and low Lp(a) levels with whole fruits (p=0.10), sodium (p=0.14), whole grains (p=0.15). There were no significant relationships between food type and high or low Lp(a) levels. HEI, Healthy Eating Index; Lp(a), Lipoprotein(a). Alpha significance set at p<0.05.

## DISCUSSION

The current study examined the effects of a free-living diet on Lp(a) plasma levels and isoforms. Since our sample was small, we first confirmed positive relationships between plasma apoB100 levels and total cholesterol (R = 0.41, p = 0.03), and LDL cholesterol (R = 0.52, p = 0.005) validating the relationship of apoB with lipids. Additionally, we found positive relationships between plasma apoB100 and dietary total fat as well as or SFA, as seen in larger cohorts, validating the relationship between diet and lipoproteins in this cohort ^35^.

Plasma Lp(a) levels are strongly determined by genetics ^5-7^, and high levels of Lp(a) are a causal risk factor for ASCVD (31). Heart healthy diets and lifestyle changes are the first steps to decreasing cardiovascular risk^14^, yet these have not been shown to lower Lp(a) levels.

Our data do not support a significant correlation between Lp(a) levels and diet macronutrient content on an absolute (grams fat, carbohydrate, or protein/day) or relative basis (percent of total calories). Results from the OMNI study showed a significant increase in Lp(a) levels with different macronutrient rich controlled diets [change in Lp(a) mean from baseline (p-value): carbohydrate + 3.2 (<0.001); protein + 4.7 (<0.001); unsaturated fat + 2.1(<0.001)] ^17^. However, this study did not control for *wIS* and race as the current study.

Current Dietary Guidelines for Americans (2020-2025) recommend consuming less than 10% of daily calories from saturated fat ^36^. The current study supports previous findings that high SFA content is linked to lower levels of Lp(a) ^15, 16, 18^. Using the average and standard deviation of SFA intake observed in our cohort (9.2 ± 4.9% Cal) and our linear regression model which includes *wIS* and SRRE, we estimate that an individual with an Lp(a) level of 100 nmol/L in whom SFA intake increases, for example, from 4% to 9%, will experience a decrease in their Lp(a) level to 70.13 nmol/L. This data can be compared to controlled studies such as the Delta study. In the latter, three different controlled diet compositions were investigated (Average American Diet (AAD) (37% Fat, 16% SFA, 14%MUFA, 7% PUFA), Step-1 Diet (30% Fat, 9% SFA, 14%MUFA, 7% PUFA), and Low Saturated Fat Diet (Low Sat) (26% Fat, 5% SFA, 14%MUFA, 7% PUFA)) and Lp(a) levels decreased as the percent of SFA of total calories was increased [(AAD: %kcal SFA=15.0±0.4, Lp(a)=15.5±1.8 mg/dL); (Step-1: %kcal SFA=9.0±0.1, Lp(a)=17.0±1.8 mg/dL); (Low Sat: %kcal SFA=6.1±0.5, Lp(a)=18.2±1.9 mg/dL)] ^18^. Two additional studies, one by Shin et al. observed increases similar to those in Delta in Lp(a) levels as participants switched from a high fat/low carb diet (40% Fat, 13% SFA, 11% MUFA, 13.8% PUFA, 3.4% trans-fat, 45% Carbohydrate, 15% Protein) to a low fat/high carb diet (20% Fat, 4.9% SFA, 9.9% MUFA, 5.1% PUFA, 2.4% trans-fat, 65% Carbohydrate, 15% Protein) diet, from an Lp(a) level of 8.91 (IQR 3.41 – 34.6) mg/dL to 11.47 (IQR 3.84 – 38.78) mg/L, respectively. Silaste et al. examined how dietary fat and vegetable consumption affect lipid levels. Using baseline and two diets [baseline (36±6 percent of total energy intake (E%) from Fat, 15±3 E% SFA, 14±3 E% MUFA, 6±1 E% PUFA, 46±7 E% Carbohydrate, 17±2 E% Protein), low fat with low vegetable (LFLV) consumption (31 E% Fat, 11 E% SFA, 13 E% MUFA, 7 E% PUFA, 49 E% Carbohydrate, 20 E% Protein) and low fat with high vegetable (LFHV) consumption (31 E% Fat, 9.5 E% SFA, 11 E% MUFA, 9.5 E% PUFA, 50 E% Carbohydrate, 20 E% Protein) ], the authors found that Lp(a) levels increased by 7% from baseline to LFLV diet and 9% from baseline to LFHV diet. More recently, a study by Ebbeling et al., showed that Lp(a) levels went down the significantly (by -14.7% when subjects consumed a low carbohydrate, high fat diet (60% of total energy from Fat, 21% SFA, 25% MUFA, 11% PUFA, 20% Carbohydrate, 20% Protein) ^37^ compared to moderate-carbohydrate diet (40% of total energy from Fat, 14% SFA, 16% MUFA, 9% PUFA, 40% Carbohydrate, 20% Protein) and high-carbohydrate diet (20% of total energy from Fat, 7% SFA, 8% MUFA, 5% PUFA, 60% Carbohydrate, 20% Protein), where Lp(a) decreased by -2.1% and increased by 0.2% without significance, respectively. A similar observation was recently described in the GET-READI, randomized crossover feeding study. In this study in African Americans, participants either consumed the American Diet with 16% SFA or Dietary Approaches to Stop Hypertension (DASH) diet with 6% SFA, for 5 weeks. Lp(a) Levels were 44mg/dl on the 16% SFA diet and 58 mg/dL with the 6% SFA diet ^38^. However, another study, where participants consumed frozen plant-based meals for 5 weeks during Lent (SFA 4.7% Kcal), researchers observed a significant reduction in Lp(a) by 10% (from 56 to 51mg/dL) ^39^. The differences in the findings reported in the latter study could be due to presence of hypertension and diabetes in the study subjects. The previous studies reported findings in otherwise healthy populations.

We see a much higher effect size (by a factor of 6 to 8) compared to the Delta study and the other two crossover studies (Shin, Silaste^15, 16^), Supplemental Figure 3. One reason can be our small cohort and the cross-sectional nature of the study. The possibility of one or two outliers influencing the regression coefficient in a cohort of 28 fitted with two continuous variables (wIS, SFA%) and three SRRE categories is real. When we looked further into intercorrelations among the predictor variables, we found that SFA% was negatively correlated with wIS and was higher in Blacks compared to the other two SRRE groups. With our current knowledge, these relationships have no biological basis and could have arisen by chance. Since the SFA% was significantly correlated with Lp(a) level only in the presence of wIS and SRRE, both correlated separately with SFA%, we would like to be conservative and conclude that the effect of SFA% on Lp(a) level is negative, while the magnitude of the effect may be overestimated due to the small cohort. The three crossover studies (Shin, Silaste, Ginsberg ^15, 16, 18^) do not have this concern since each subject was studied at different SFA% levels, and so each subject served as their own control.

In the current study we were able to include apo(a) isoform size and SRRE, both of which are known to affect Lp(a) levels. Considering these, improved our negative associations of Lp(a) levels with total SFA and palmitic and stearic acids. Neither our study nor previous studies have performed metabolic studies that could help elucidate the mechanisms that are regulating these reported associations. The lower Lp(a) levels with diets high in SFA could be due to decreased production of Lp(a) particles.

A possible mechanism for this relationship could be that high SFA intake could change the fatty acid profile in the phospholipid membrane of apoB100-containing particles to create a less favorable arrangement for apo(a) to bind and hence decrease production, resulting in a lower Lp(a) concentration. There are no data reported for understanding the effects of macronutrients on synthesis or production of Lp(a) particles.

Another possible mechanism, as mentioned by Enkhmaa et al., is that lower SFA diets could reduce the clearance of Lp(a) particles via the LDL receptor ^40^. This could be attributable to increased competition with other apoB100 containing particles, including LDL. A recent study from our group showed that both production and clearance of Lp(a) and isoforms regulate its level ^32^. Therefore, we speculate that diet composition may be regulating Lp(a) levels through combined mechanisms.

Although a small study, our calculated diet quality based on HEI analysis is similar to those reported in larger cohort studies ^34^. The macronutrient distribution is similar to the findings reported by the National Health and Nutrition Examination Survey (NHANES) data (2017-2018), where the percent of calories from carbohydrate, protein, at, and saturated fatty acids were 47.3%, 16%, 34.8%, and 11.3%, respectively ^34^. The HEI score provides a way to assess diet quality, and we hypothesized that a higher HEI score (better overall diet quality) would correlate with a lower level of Lp(a). However, we found no significant relationship between HEI and Lp(a) levels including saturated fat. Importantly, when stratified on Lp(a) level (≤ or > 100nmol/L) and high (>100nmol/L), we observed a statistically significant relationship with saturated fat. The low Lp(a) group had a lower score for saturated fat (58%) [compared to high (87%)], meaning they consumed more saturated fat and thus received a lower score when HEI was calculated ^25^. This data supports our overall findings of our study.

Our study has several limitations: (1) We had a limited sample size as the data were obtained from subjects previously enrolled in small studies that examine lipid metabolism in humans, (2) The study was observational in nature and, (3) the data based on subjects who completed 24-hour hour food records. There are known limitations to 24-hour dietary intake data such as recall bias, which may be skewed based on the subject’s desire to express their intake to the recorder. It is possible that participants under or over-reported various foods or left out stereotypically undesirable foods entirely. However, these subjects were not enrolled for a diet intervention study. Photographic or meal-logging systems could have been used to help minimize this bias, however these dietary assessment tools may impart additional bias as participants may consciously or subconsciously change their intake whenever diet information is collected. Additionally, only one recall per participant was analyzed for this study. Future studies should take a more comprehensive approach to examine typical intake by issuing multiple food records (on weekdays and weekends) and throughout the year to account for seasonable variability in food intake.

Despite these limitations, the study population did have similar SFA intake compared to larger controlled randomized diet studies that have similar findings.

The National Cholesterol Education Program Adult Treatment Panel (NCEP ATP) III and the American Heart Association (AHA) recommend therapeutic lifestyle changes (TLC) as the primary treatment for lowering LDL-C ^12, 41, 42^ and improving cardiovascular health. High Lp(a) levels are causal for ASCVD, hence, understanding the exact mechanisms that regulate diet composition effects on Lp(a) are important. Specifically, understanding how diet affects Lp(a) lipid composition and synthesis may be a clinical important area of study.

Our findings support that increased dietary saturated fat is associated with low Lp(a) levels. Together with the growing field of nutrigenomics ^43^, it is possible that individualized diet recommendations can be tailored to address a patient’s ASCVD risk profile and hopefully future studies will focus on what is best for individuals with high levels of Lp(a).

## Data Availability

All the data are presented within the article and supplementary material. Additional data queries are available via direct contact with corresponding author.

